# Associations between Acute Treatments for Spinal Cord Strokes and Functional Outcomes

**DOI:** 10.64898/2026.03.24.26349240

**Authors:** Trevor Glenn, Philippe Bilodeau, Ahya Ali, Shamik Bhattacharyya

## Abstract

**Background:** Acute treatments for patients with spinal cord strokes (SCS), including lumbar drain, blood pressure augmentation, corticosteroids, antiplatelets, and anticoagulants, are largely extrapolated from literature on cerebral infarcts or based on suspected SCS physiology. This study adds to the knowledge of symptomatology and management of SCS.

**Methods:** This retrospective cohort study included patients from one medical system from 2000-2025. Multivariate ordinal logistic regressions were performed to evaluate associations of SCS treatments with the primary outcome of ambulatory status (independently ambulatory, ambulatory with assistance, non-ambulatory) at first follow-up, as well as secondary outcomes of modified Rankin Scale (mRS) and modified Japanese Orthopedic Association (mJOA) scores. SCS severity by American Spinal Injury Association impairment scale (AIS) with grade A as the comparator, age, sex, and whether SCS was spontaneous/periprocedural were covariates. Odds ratios (OR) greater than 1 were associated with better ambulatory status, lower mRS, and higher mJOA.

**Results:** 130 SCS patients were included. Median age at SCS onset was 62 years, 42% were female, and 39% were periprocedural. Median first follow-up was 57 days. AIS grade was A for 28%, B for 25%, C for 28%, and D for 26%. SCS severity had significant associations with outcomes. For ambulatory status, AIS B OR 2.78, 95% CI 1.03-7.69, p-value 0.045; AIS C OR 16.7, 95% CI 5.56-50.0, p-value <0.01; AIS D OR 125, 95% CI 33.3-500, p-value <0.01. Corticosteroids were associated with improved ambulatory status and mJOA at follow-up (OR 2.38, 95% CI 1.15-5, p-value 0.023 and OR 2.27, 95% CI 1.09-4.76, p-value 0.030, respectively). No treatment had a significant association with mRS.

**Conclusion:** Initial SCS severity had the strongest association with outcomes. Corticosteroids were associated with a better ambulatory status and mJOA. This study can help guide clinician management of patients with SCS.

## Introduction

Spinal cord strokes (SCS) represent around 1.2% of ischemic strokes; however, this may be an underestimate since as many as 14-16% of patients referred for transverse myelitis have SCS^1–3^. SCS occurs spontaneously or post-procedurally and manifests as a variety of symptoms, such as acute onset weakness, numbness, bowel/bladder dysfunction, or dysautonomia.

Evidence-based therapies for the acute treatment of SCS are needed. Acute treatments include CSF drainage by lumbar drain, blood pressure augmentation, corticosteroids, antiplatelet agents, and anticoagulants. These therapies are mostly extrapolated from literature on cerebral infarcts or suspected SCS physiology^4–7^.

CSF drainage by lumbar drain placement is more unique to SCS and is hypothesized to improve spinal cord perfusion by lowering the pressure in the spinal subarachnoid space. Lumbar drain placement is associated with potential complications of subdural hematoma and central nervous system infection^8^. Placement of lumbar drains preoperatively for open thoracoabdominal aneurysm repair or endovascular aneurysm repair helps decrease the risk of SCS, but the utility of lumbar drains *after* SCS is less certain, and there is also a lack of data regarding the role for lumbar drains in spontaneous SCS ^9,10^. One small case series of three patients with SCS (either periprocedural or secondary to type B aortic dissection) did report improvement in motor function after lumbar drain placement, although additional data with larger patient populations would be benficial^11^.

Similarly, data are lacking for other interventions used for SCS such as corticosteroids and blood pressure augmentation. There are studies and guidelines more broadly from spinal cord injury (SCI) literature that explore the utility of these interventions for treatment of SCI.

The AO Spine 2017 practice guidelines recommend treatment with 24 hours of methylprednisolone starting within 8 hours of spinal cord injury^12^. Other meta-analyses of observational data, however, have not found a benefit of methylprednisolone for SCI when used within 8 hours of injury, and some have documented increased risk of gastrointestinal bleeding or respiratory illness with use of high-dose steroids^13,14^. Guidelines recommend blood pressure augmentation for management of acute spinal cord injury by elevating the mean arterial pressure (MAP) to greater than 85 mmHg for 7 days post spinal cord injury; however, data from randomized trials for spontaneous SCS are lacking^15^. A prior review evaluated nine retrospective studies and two prospective cohort studies investigating blood pressure management in spinal cord injury^16^. The data were mixed with some studies suggesting improved outcomes with blood pressure augmentation, some that reported worse neurologic outcomes, and some that found no difference^17–21^. A recent RCT that compared augmented and conventional blood pressure goals in patients with spinal cord injury found no differences in sensory or motor scores between the two groups and higher rates of complications in the augmented blood pressure arm^22^. While these studies were done in cases of SCI rather than SCS, the treatments evaluated in these studies are often utilized in patients with SCS. To address these gaps in evidence, we evaluated data from our center to assess the impact of SCS acute treatments on ambulatory and functional outcomes.

## Methods

### Patient Selection and Measurement of Outcomes

The study retrospectively evaluated 130 patients diagnosed with SCS between 2000 and 2024 in the Mass General Brigham health system, which consists of 16 academic and community hospitals. The institutional review board at Mass General Brigham approved the study; no informed consent was required. Electronic medical records were searched for more than two instances of the ICD10 diagnostic code G95.11 (acute infarction of the spinal cord). 211 patients met this criterion. Medical records were then examined for a clinical diagnosis of SCS that was sustained on follow-up and continued assessment. 81 patients were excluded due to alternative final diagnoses, and 130 patients were included. Demographics, vascular risk factors, clinical presentations, treatments, and outcomes at discharge and first outpatient follow-up were collected from the electronic medical record. All authors had full access to all the data in the study. The STROBE reporting guidelines were utilized for this retrospective cohort study.

The Zalewski categorization system for spinal cord strokes, which takes into account the time course of symptoms, whether it was spontaneous or periprocedural, MRI findings, and CSF findings, was used to classify types of SCS (as definite, probable, or possible)^23^. The American Spinal Injury Association (ASIA) impairment scale (AIS)^24^ was used to classify SCS severity.

The modified Rankin Scale (mRS)^25^ and modified Japanese Orthopedic Association scale (mJOA)^26^, which takes into account motor function of the upper extremities, motor function of the lower extremities, sensory function, and sphincter function (particularly regarding ability to urinate), were assessed retrospectively for clinical outcomes after SCS.

### Outcomes

The primary study outcome was ambulatory status at the first follow-up visit classified as (1) independently ambulatory, (2) ambulatory with assistance, or (3) non-ambulatory. Ambulatory status was stratified by acute treatment for SCS (antiplatelet, anticoagulant, blood pressure augmentation, corticosteroid, and lumbar drain), which were not mutually exclusive. Secondary outcomes were mRS and mJOA scores at the first follow-up visit. The first follow-up visit was used because of the variable length of clinical follow-up, and many patients only had a single follow-up visit. Seven patients (5%), however, did not have outpatient follow-up information available in our medical record system. For these patients, the last observation for ambulatory status, mRS, and mJOA were carried forward so that these patients were still included in the outcome analyses.

### Statistical Analysis

Statistical analyses were run using R Statistical Software (v4.1.2; R Core Team, 2021). Given that the continuous data were not normally distributed, median values and interquartile ranges (IQR) were utilized. Fischer’s Exact and Mann-Whitney U tests were used to determine statistically significant differences between groups that did or did not receive the treatments of interest. Multivariate ordinal logistic regressions were performed to determine how acute SCS treatments were associated with the primary outcome of ambulatory status at first follow-up (adjusting for multiple comparisons), as well as for the secondary outcomes of mRS and mJOA scores. SCS severity by AIS, age, sex, and whether the SCS was spontaneous or periprocedural were covariates in the models. Statistical significance was set to p < 0.05.

## Results

### Demographic Information and Characteristics

Table 1 shows demographic information for the 130 patients at the time of SCS onset. The median age of onset was 62 years (IQR 45, 70), and 42% were female. The median BMI was 26 (IQR 23-31), 45% had a history of hypertension, 23% had hyperlipidemia, 23% had coronary artery disease, 20% had diabetes mellitus, 18% had atrial fibrillation, and 3% had a prior cerebral stroke.

**Table 1.** Demographic information.

| **Characteristic** | **N=130\*** |
| --- | --- |
| Age | 66 (52, 75) |
| Sex |  |
| Female | 54 (42%) |
| Male | 76 (58%) |
| Race |  |
| Black or African American | 10 (7.7%) |
| White | 117 (90%) |
| Other | 3 (2.3%) |
| Ethnicity |  |
| Hispanic or Latino | 6 (4.6%) |
| Not Hispanic or Latino | 124 (95%) |
| BMI | 26 (23, 21) |
| Smoking History |  |
| Current | 20 (15%) |
| Former | 47 (36%) |
| Never | 63 (48%) |
| History of Significant Alcohol Use | 9 (6.9%) |
| Systemic Cancer | 11 (8.5%) |
| Hypertension | 59 (45%) |
| Diabetes | 26 (20%) |
| Hyperlipidemia | 30 (23%) |
| Prior Stroke | 4 (3.1%) |
| Coronary Artery Disease | 30 (23%) |
| Atrial Fibrillation | 23 (18%) |

\* Median (IQR); n (%)

Table 2 contains information about SCS presentation, management, and disposition. 39% of SCSs were periprocedural while 61% were spontaneous. Of the periprocedural cases, 48% were open procedures and 52% were endovascular. There were 19 different types of procedures associated with SCS (Supplementary Table 1). Aortic aneurysm repairs were the most common procedure (30 patients), followed by cervical and/or thoracic laminectomies (3 patients), aortic dissection repairs (2 patients), and initiation of extracorporeal membrane oxygenation cannulas (2 patients). The median time from symptom onset to symptom nadir was less than an hour (IQR 0, 2). 38% reported pain with the onset of symptoms. AIS grade at nadir of symptoms was A for 28%, B for 25%, C for 28%, and D for 26%. 12% of patients had spinal cord strokes diagnosed acutely and at follow-up but had symptom progression from onset to nadir lasting longer than 12 hours, which did not meet Zalewski criteria (although there were imaging findings or other evidence suggestive of SCS). CSF studies were obtained acutely in 37 (28.5%) of cases. 4 of the 37 (10.8%) patients had a white blood cell count greater than five cells/microliter (the highest was 24 cells/microliter). 3 of 4 cases had a neutrophilic predominance – the other case was lymphocytic. Oligoclonal bands were not detected in the CSF of the patients with elevated white blood cell counts. The median CSF protein was 36.5 mg/dL (IQR 31.8, 42.5).

**Table 2.** Spinal cord stroke (SCS) classification, management, and disposition.

| Characteristic | N=130* |
| --- | --- |
| SCS Classification |  |
| Definite Periprocedural | 41 (32%) |
| Definite Spontaneous | 40 (31%) |
| Probable Periprocedural | 7 (5.4%) |
| Probable Spontaneous | 21 (16%) |
| Possible Spontaneous | 6 (4.6%) |
| Did Not Meet Criteria | 15 (12%) |
| Age of Onset | 62 (45, 70) |
| Time to Symptom Nadir (Hours) | 0.03 (0, 2) |
| Pain with Onset of Symptoms | 49 (38%) |
| AIS<sup>†</sup> Scale at Nadir |  |
| A (no motor, no sensory) | 28 (22%) |
| B (no motor) | 32 (25%) |
| C (50% of muscles less than 3) | 36 (28%) |
| D (50% of muscles more than 3) | 34 (26%) |
| Etiology |  |
| Perioperative | 51 (39%) |
| Spontaneous | 79 (61%) |
| Procedure Type |  |
| Open | 24 (48%) |
| Endovascular | 26 (52%) |
| Intervention for SCS |  |
| Blood Pressure Augmentation | 54 (42%) |
| Lumbar Drain | 33 (25%) |
| Antiplatelet Agent | 74 (57%) |
| Anticoagulation | 21 (16%) |
| Corticosteroids | 33 (25%) |
| Length of Hospitalization | 12 (8, 25) |
| Discharge Disposition |  |
| Home | 17 (13%) |
| Inpatient Rehab | 98 (77%) |
| Skilled Nursing Facility | 2 (1.6%) |
| Deceased | 7 (5.4%) |
| Time to First Follow-Up (Days) | 57 (30, 131) |

\*Median (IQR); n (%).
<sup>†</sup>American Spinal Injury Association impairment scale

### Treatments

Patients were treated with an antiplatelet agent in 57% of cases, blood pressure augmentation in 42%, lumbar drain in 25%, corticosteroids in 25%, and anticoagulation in 16%. The median time from SCS onset to lumbar drain placement was 16 hours (IQR 4, 29) with a median duration of lumbar drain placement of 90 hours (IQR 61, 114). The median time from SCS onset to initiation of steroids was 25 hours (IQR 7, 72.5) with a median duration of 49 hours (IQR 48, 96). Corticosteroid dose and duration varied widely, such as three days of 1 g of intravenous methylprednisolone to 10 mg of dexamethasone once to 4 mg of dexamethasone every 4 hours for four days. The median time from SCS onset to initiation of blood pressure augmentation was 12.25 hours (IQR 4.63, 26.75) with a median time of augmentation of 66.5 hours (IQR 43.5, 93.75) (Supplementary Figure 1). A mean arterial pressure target was more frequently chosen than a systolic blood pressure target (83% compared to 17% of the time, respectively). The median mean arterial pressure target was greater than 85 mmHg (IQR 80, 85). Supplementary Table 2 shows differences in demographics, SCS etiology, and AIS grade between groups that did or did not receive the various acute treatments of interest. Notably, of those who underwent lumbar drain placement, 72.7% had a perioperative SCS. Of those who received corticosteroids, 87.9% had a spontaneous SCS.

### Outcomes

The median length of hospitalization was 12 days (IQR 8, 25) (Table 2). At discharge, 18 (14%) were ambulatory without assistance, 26 (20%) were ambulatory with assistance, and 86 (66%) were non-ambulatory. 13% were discharged home, 77% to an inpatient rehabilitation center, 1.6% to a skilled nursing facility, and 5% were deceased. The median first follow-up time was 57 days (IQR 30, 131), and 29 (22%) were ambulatory without assistance, 52 (40%) were ambulatory with assistance, and 49 (38%) were non-ambulatory (Supplementary Figure 2). At discharge, 16 had an mRS of 1 (12%), 3 patients had an mRS of 2 (2%), 22 had an mRS of 3 (17%), 26 had an mRS of 4 (20%), 52 had an mRS of 5 (40%), and 11 had an mRS of 6 (8%). At follow-up, 3 patients had an mRS of 0 (3%), 16 had an mRS of 1 (14%), 21 had an mRS of 2 (19%), 28 had an mRS of 3 (25%), 23 had an mRS of 4 (20%), and 21 had an mRS of 5 (19%).

At discharge, 15 (11%) had a mild myelopathy (mJOA 15 to 17), 22 (17%) had a moderate myelopathy (mJOA 12 to 14), and 93 (72%) had a severe myelopathy (mJOA 0 to 11). At follow-up, 22 (17%) had a mild myelopathy, 33 (25%) had a moderate myelopathy, and 75 (58%) had a severe myelopathy. On average, patients had better ambulatory status, a lower mRS and a higher mJOA over time. The median mRS scores at nadir, discharge, and follow-up were 4, 4, and 3, respectively. The median mJOA scores at nadir, discharge, and follow-up were 9, 10, and 12, respectively.

Figure 1A shows the association between acute treatments and ambulatory status at first follow-up, adjusted for age, sex, etiology, initial severity, and other concurrent treatments. An odds ratio (OR) greater than 1 is associated with better ambulatory status. Compared to AIS grade A, better grades were associated with better ambulatory status at first follow-up (AIS B OR 2.78, 95% CI 1.03-7.69, p-value 0.045; AIS C OR 16.7, 95% CI 5.56-50.0, p-value <0.01; AIS D OR 125, 95% CI 33.3-500, p-value <0.01). The only acute intervention associated with improved ambulatory status at first follow-up was corticosteroid use (OR 2.38, 95% CI 1.15-5, p-value 0.023). Figures 1B and 1C show secondary outcomes of mRS and mJOA at first follow-up. An OR greater than 1 is associated with a lower mRS (better outcome). An OR of greater than 1 is associated with a higher mJOA (better outcome). Similarly, compared to AIS grade A, better AIS grades were associated with lower mRS (AIS C OR 7.69, 95% CI 2.85-20.0, p-value <0.01; AIS D OR 100, 95% CI 25.0-250, p-value <0.01) and higher mJOA (AIS C OR 12.5, 95% CI 4.54- 33.3, p-value <0.01; AIS D OR 100, 95% CI 24.4-250, p-value <0.01). Corticosteroid use was also associated with a higher mJOA (OR 2.27, 95% CI 1.09-4.76, p-value 0.030). No acute treatments were associated with improved mRS at follow-up.

**Figure 1.**
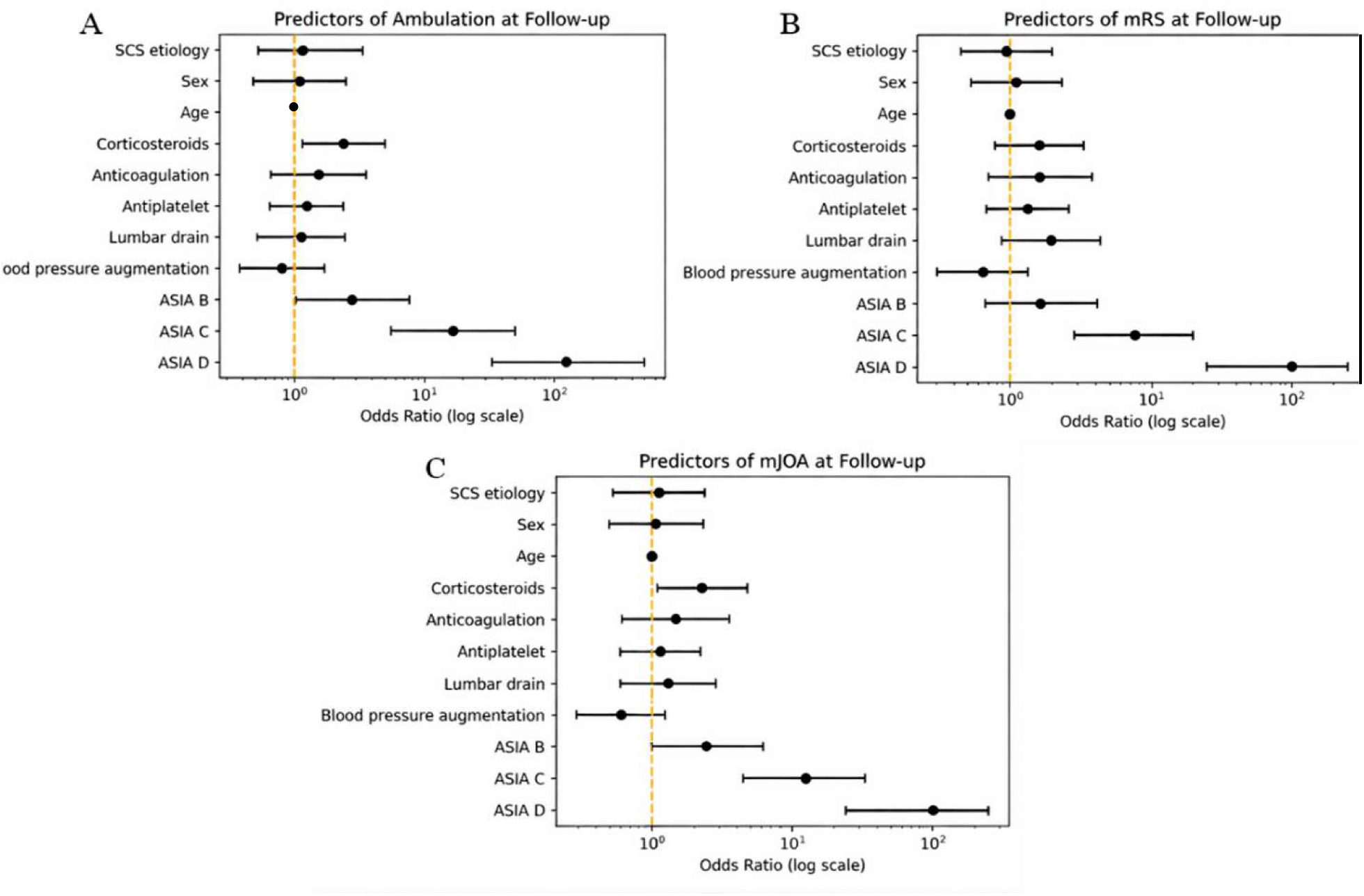
Association between covariates and acute treatments with outcomes of A. Ambulatory status B. Modified Rankin Scale (mRS), and C. Modified Japanese Orthopaedic Association (mJOA) at first follow-up.

## Discussion

Patient presentations of spinal cord strokes can be diverse, and etiologies of the infarct can vary, with different causes potentially benefiting from different management strategies. There are few guidelines and consensus statements regarding SCS diagnosis, classification, and management. Data are further limited because some studies only evaluate SCS of a specific etiology. SCS can be periprocedural or spontaneous, and potential management differences among etiologies are poorly delineated. SCS can be debilitating, and prior studies have cited a variable but overall high degree of functional dependence after SCS^27,28^. Specifically, one prior study found that 2 out of 27 (1%) patients with a periprocedural SCS and 19 out of 29 (66%) patients with spontaneous SCS regained the ability to walk^29^. Another study showed that around 40% of individuals who were wheelchair bound at the time of discharge could walk at follow-up^30^. A meta-analysis of 440 patients reported 71% of patients were ambulatory with or without a walking aid at follow-up (although follow-up was sometimes many years after the SCS)^31^.

Given the rarity, heterogeneity, and significant morbidity associated with SCS, this study aimed to add to the growing knowledge of symptomatology, management, and outcomes of patients with cord infarcts. We describe the clinical presentations of 130 patients diagnosed with SCS, the treatments utilized, and the factors associated with ambulatory status, lower mRS, and improved mJOA to provide more information to help guide physicians managing patients with this condition. While 12% of our cases did not meet diagnostic criteria (as put forth by Zalewski et al. 2018) due to prolonged time from symptom onset to nadir, only 77% of the cases in that cohort had reached nadir by 12 hours (the others had a more stuttering course)^23^. While those authors had recognized that some autopsy-proven cases of SCS evolved over more than 12 hours, they had chosen the cutoff of 12 hours to help delineate SCS from other spinal cord syndromes that typically evolve over a longer period of time (sacrificing sensitivity for specificity). However, time from symptom onset to nadir, while typically rapid, can be more prolonged in SCS than in cerebral strokes.

The most significant predictor of mRS and mJOA in our cohort was the initial severity of SCS, similar to what has been previously described^30^. The use of corticosteroids was associated with better ambulatory status and a better mJOA score at follow-up. While there is no evidence for a role for corticosteroids in the treatment of cerebral strokes, the different mechanisms of SCS compared to cerebral stroke may lead to local inflammatory aspects in SCS pathophysiology that could benefit from anti-inflammatory effects of corticosteroids, although further studies with larger populations are necessary to better evaluate the utility of corticosteroids and other treatments for SCS. Of note, the majority of patients (87.9% who received corticosteroids had spontaneous rather than periprocedural SCS; thus, the possible correlation between steroids and outcomes should be interpreted with caution, particularly regarding patients with periprocedural SCS. Given the imbalance in the use of corticosteroids, there may be residual confounding relating to the nature of the infarct (whether it was spontaneous or periprocedural) that may be impacting the association between steroids and ambulatory ability (even though this was held as a covariate in the model). Of the other therapies more unique to SCS, blood pressure augmentation and lumbar drains did not have a statistically significant association with ambulatory status, mJOA, or mRS in this cohort. While these interventions did not reach statistical significance, many patients in our cohort improved over time: patients’ ambulatory status and median mRS and mJOA scores improved from discharge to follow-up.

This study has its limitations. Not all of the cases were able to be classified as definitive spontaneously or perioperative SCS; thus, it is possible that some of the patients were incorrectly diagnosed with SCS. Some patients did not have primary imaging data within our system; therefore, we relied on outside radiology reports. The mJOA was initially intended for cervical myelopathy but was extended to patients without cervical cord involvement in this study (we did not restrict the mJOA to patients with cervical involvement, as that would have further decreased our sample size/limited the study’s power). The power for some aspects of the study was limited by the low number of certain interventions, such as anticoagulation (utilized in 16% of patients) and lumbar drains or corticosteroids (used in 25% of patients). Additionally, certain interventions were utilized more in certain populations (patients who received corticosteroids were more likely to have spontaneous SCS and those who received a lumbar drain were more likely to have periprocedural SCS), which impacts the generalizability of the results. Corticosteroid dose and duration were not standardized and instead varied greatly in this population. Given the low number of certain interventions, we did not separate cases based on etiology of SCS (either spontaneous or periprocedural) but included it as a covariate in our model; however, there may be differences in optimal acute management of SCS based on underlying etiology that we were unable to explore with this data set. Additionally, the median follow-up time was somewhat short at 57 days, and there may have been benefits from certain interventions that would have only become apparent further out from the SCS.

The data from this cohort show that the factor most predictive of ambulatory status, mRS, and mJOA at first outpatient follow-up is initial SCS severity as graded by the ASIA scale. The use of steroids was also associated with having a better ambulatory status and mJOA at follow-up. Other interventions (blood pressure augmentation, placement of a lumbar drain, use of antiplatelet or anticoagulation) did not have a statistically significant association with ambulatory status, mRS, or mJOA. On average, patients had improvement in their mRS and mJOA scores from discharge to follow-up. More data are needed to help improve treatment recommendations and, thus, outcomes in patients with SCS.

## Non-standard Abbreviations and Acronyms

SCS: spinal cord stroke
SCI: spinal cord injury
ASIA: American Spinal Injury Association
AIS: ASIA impairment scale
mRS: modified Rankin scale
mJOA: modified Japanese Orthopedic Association

## Data Availability

Data could be available upon reasonable request

---

**Supplementary Figure 1.**
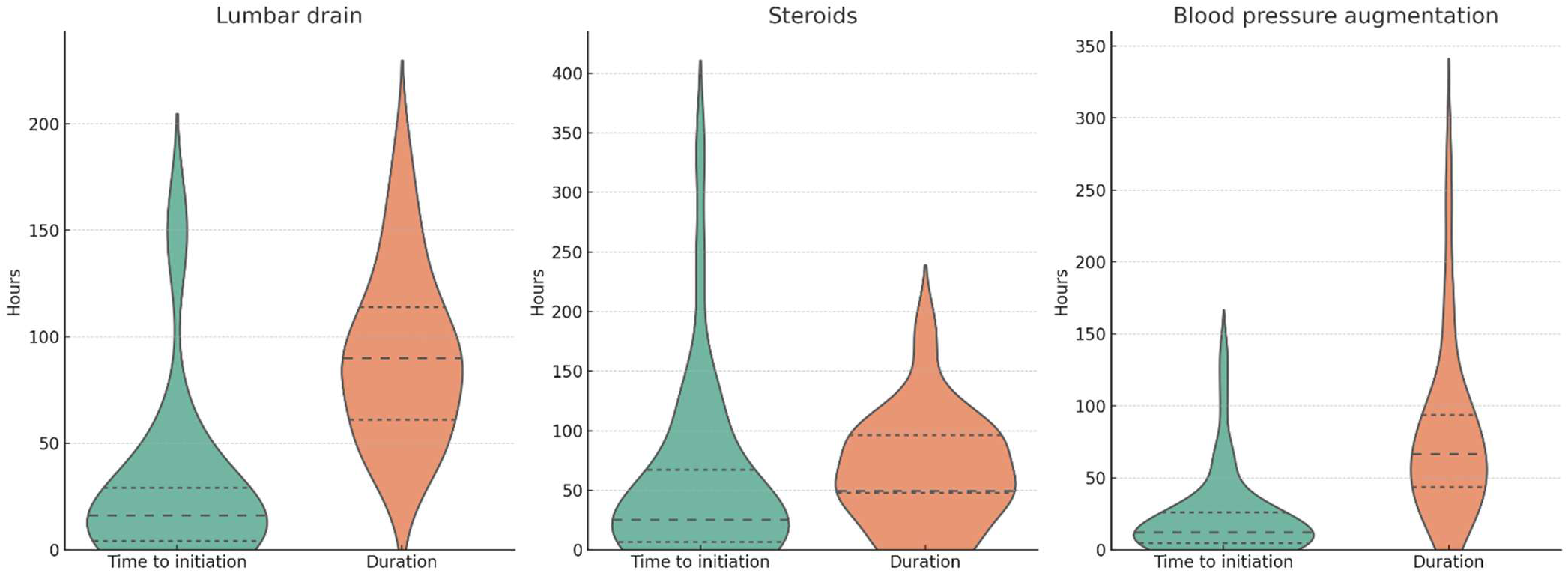
Time to initiation and duration of lumbar drain, corticosteroids, and blood pressure augmentation

**Supplementary Figure 2.**
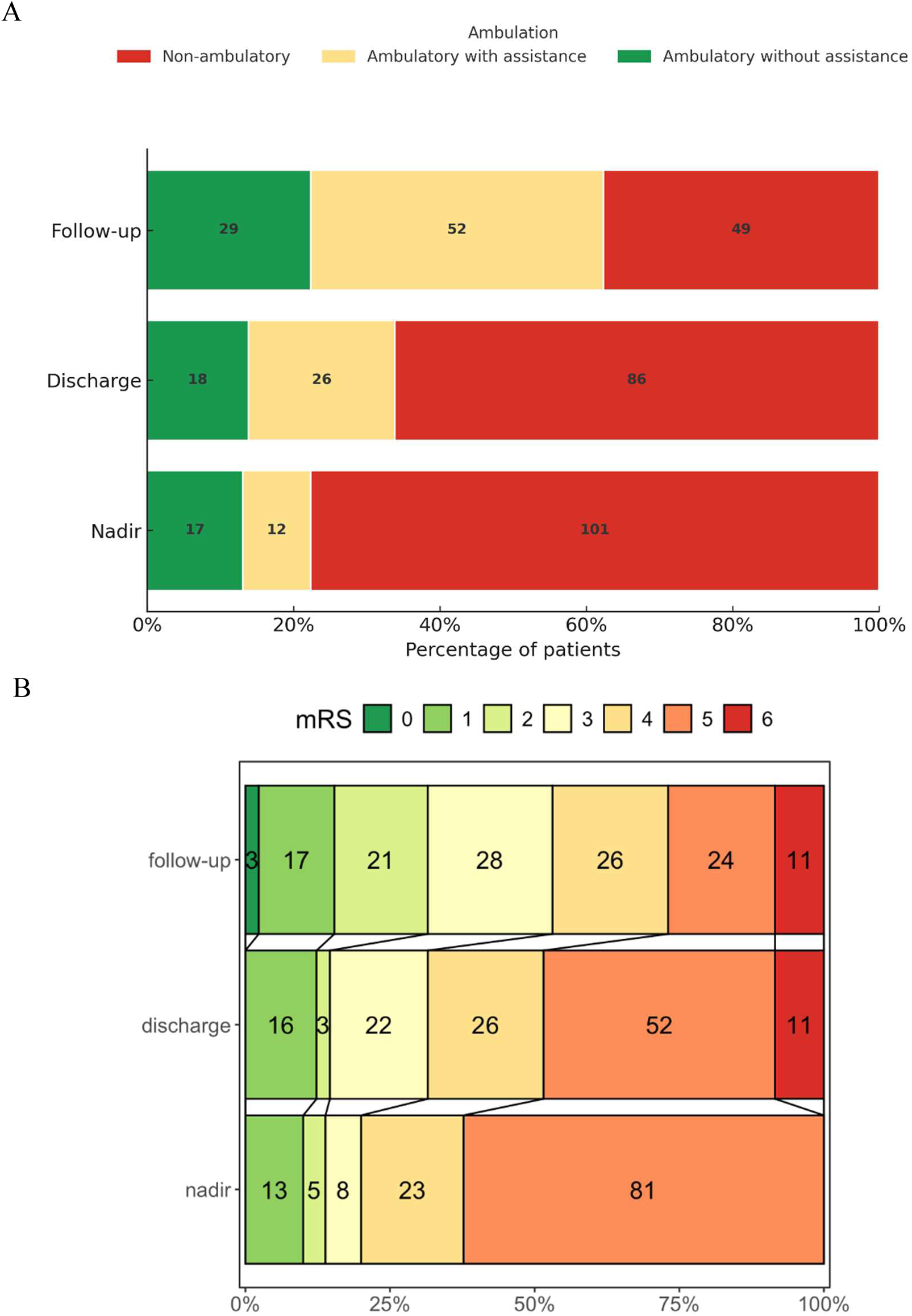
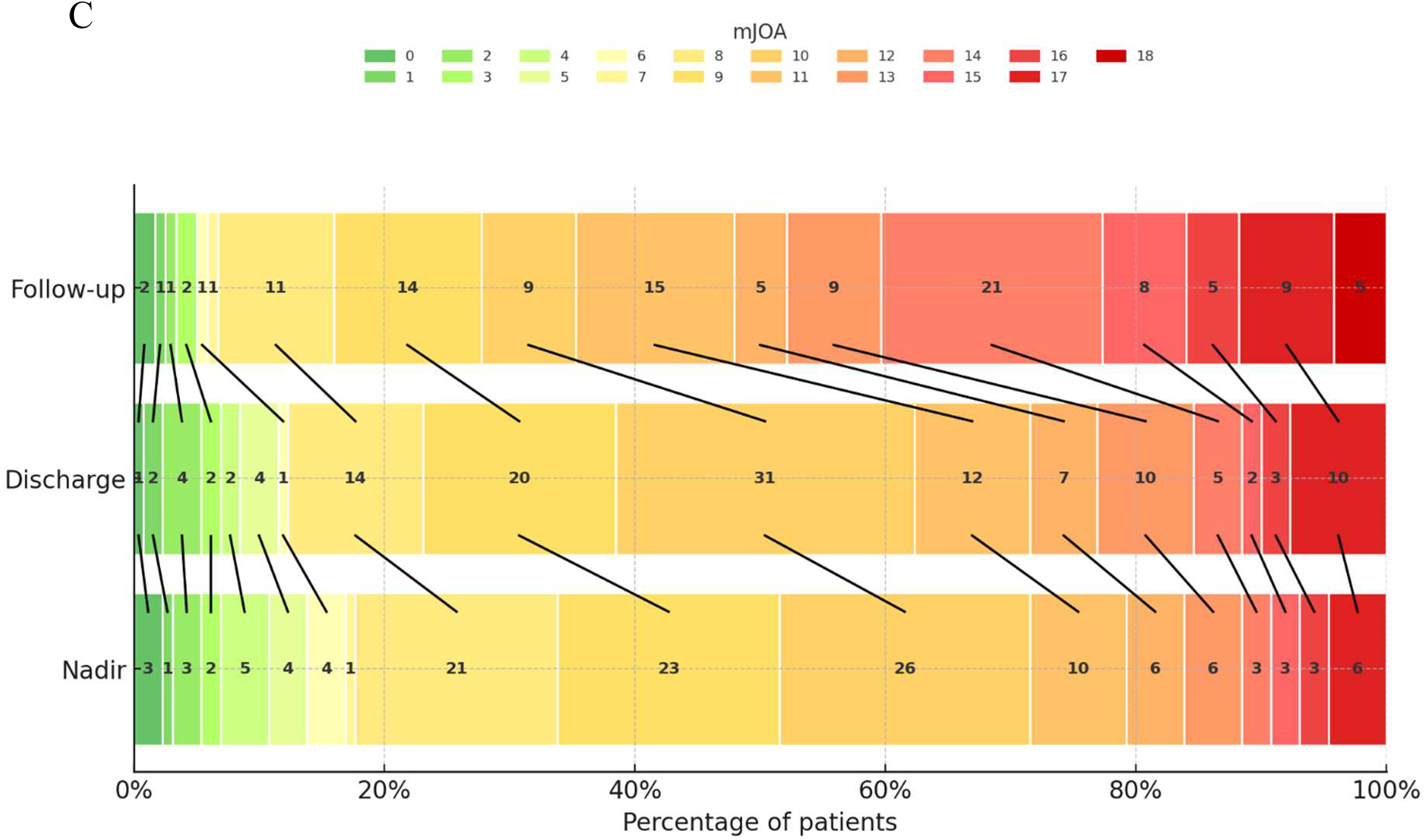
Changes in A. Ambulatory status, B. Modified Rankin Scale (mRS), and C. Modified Japanese Orthopaedic Association (mJOA) over time.

**Supplementary Table 1.** Procedure types associated with spinal cord strokes.

| **Procedure type** | **Number of cases** |
| --- | --- |
| Aortic aneurysm repair | 30 |
| Cervical and/or thoracic laminectomy and fusion | 3 |
| Aortic dissection repair | 2 |
| Extracorporeal membrane oxygenation | 2 |
| Right lower lobe lung resection | 1 |
| Superior mesenteric artery (SMA) endarterectomy and supraceliac aorta-SMA bypass | 1 |
| Excision of ruptured splenic pseudoaneurysm | 1 |
| Aortic valve replacement c/b aortic dissection | 1 |
| Simultaneous left ventricular assist device and right ventricular assist device placement | 1 |
| Digital subtraction angiography | 1 |
| Embolization of intercostal artery pseudoaneurysm rupture | 1 |
| Right cervicomedullary arteriovenous malformation embolization | 1 |
| Aortobifemoral bypass | 1 |
| Intra-arterial thrombectomy | 1 |
| Coronary angiography | 1 |
| Washout of infected L2-L5 fusion | 1 |
| Mitral valve repair c/b aortic dissection | 1 |
| Embolization of dural arteriovenous fistula | 1 |

**Supplementary Table 2.** Differences in demographics and spinal cord stroke severity between those who did and did not receive various treatments. A. Blood pressure augmentation. B. Placement of a lumbar drain. C. Use of Corticosteroids. D. Use of an antiplatelet agent. E. Use of anticoagulation.

**A. Blood pressure augmentation**
| Characteristic | Yes (n=54) | No (n=76) | p value |
| --- | --- | --- | --- |
| Female, % | 21 (38.9%) | 33 (43.4%) | 0.72 |
| Age, median (IQR) | 66 (46.5, 77.0) | 66 (52, 74) | 0.94 |
| Hypertension, n (%) | 27 (50.0%) | 32 (42.1%) | 0.376 |
| Diabetes, n (%) | 12 (22.2%) | 14 (18.4%) | 0.597 |
| Hyperlipidemia, n (%) | 14 (25.9%) | 16 (21.1%) | 0.52 |
| Coronary artery disease, n (%) | 15 (27.8%) | 15 (19.7%) | 0.287 |
| Atrial fibrillation, n (%) | 9 (16.7%) | 14 (18.4%) | 0.8 |
| Perioperative, n (%) | 29 (53.7%) | 25 (46.3%) | **0.0061** |
| AIS* nadir, median (IQR) | 2 (1, 3) | 3 (2, 4) | **0.0067** |

**B. Lumbar drain**
| Characteristic | Yes (n=33) | No (n=97) | p value |
| --- | --- | --- | --- |
| Female, % | 13 (39.4%) | 41 (42.3%) | 0.84 |
| Age, median (IQR) | 64 (52, 74) | 70 (52, 78) | 0.26 |
| Hypertension, n (%) | 20 (60.6%) | 39 (40.2%) | **0.043** |
| Diabetes, n (%) | 7 (21.2%) | 19 (19.6%) | 0.84 |
| Hyperlipidemia, n (%) | 7 (21.2%) | 23 (23.7%) | 0.77 |
| Coronary artery disease, n (%) | 12 (36.4%) | 18 (18.6%) | **0.037** |
| Atrial fibrillation, n (%) | 10 (30.3%) | 13 (13.4%) | **0.029** |
| Perioperative, n (%) | 24 (72.7%) | 28 (28.9%) | **<0.01** |
| AIS* nadir, median (IQR) | 2 (2, 2) | 2 (2, 2) | 1 |

**C. Corticosteroids**
| Characteristic | Yes (n=33) | No (n=97) | p value |
| --- | --- | --- | --- |
| Female, % | 18 (54.5%) | 36 (37.1%) | 0.1 |
| Age, median (IQR) | 62 (46.5, 77.0) | 68 (54, 77) | **0.015** |
| Hypertension, n (%) | 10 (30.3%) | 49 (50.5%) | **0.045** |
| Diabetes, n (%) | 9 (27.3%) | 17 (17.5%) | 0.23 |
| Hyperlipidemia, n (%) | 5 (15.2%) | 25 (25.8%) | 0.214 |
| Coronary artery disease, n (%) | 5 (15.2%) | 25 (25.8%) | 0.214 |
| Atrial fibrillation, n (%) | 3 (9.1%) | 20 (20.6%) | 0.136 |
| Perioperative, n (%) | 4 (12.1%) | 29 (87.9%) | **<0.001** |
| AIS* nadir, median (IQR) | 2 (2, 4) | 3 (2, 3) | 0.83 |

D. Antiplatelet
| Characteristic | Yes (n=74) | No (n=56) | p value |
| --- | --- | --- | --- |
| Female, % | 37 (50.0%) | 17 (30.4%) | **0.031** |
| Age, median (IQR) | 68 (54.5, 75.0) | 62.5 (47.8, 73.2) | 0.16 |
| Hypertension, n (%) | 36 (48.6%) | 23 (41.1%) | 0.39 |
| Diabetes, n (%) | 13 (17.6%) | 13 (23.2%) | 0.43 |
| Hyperlipidemia, n (%) | 23 (31.1%) | 7 (12.5%) | **0.013** |
| Coronary artery disease, n (%) | 22 (29.7%) | 8 (14.3%) | **0.04** |
| Atrial fibrillation, n (%) | 9 (12.2%) | 14 (25.0%) | 0.059 |
| Perioperative, n (%) | 26 (35.1%) | 48 (64.9%) | 0.28 |
| AIS* nadir, median (IQR) | 3 (2, 4) | 2 (1.75, 3) | 0.082 |

E. Anticoagulant
| Characteristic | Yes (n=21) | No (n=109) | p value |
| --- | --- | --- | --- |
| Female, % | 8 (38.1%) | 46 (42.2%) | 0.81 |
| Age, median (IQR) | 70 (57, 76) | 65 (52, 75) | 0.47 |
| Hypertension, n (%) | 11 (52.4%) | 48 (44.0%) | 0.486 |
| Diabetes, n (%) | 5 (23.8%) | 21 (19.3%) | 0.638 |
| Hyperlipidemia, n (%) | 6 (28.6%) | 24 (22.0%) | 0.518 |
| Coronary artery disease, n (%) | 6 (28.6%) | 24 (22.0%) | 0.518 |
| Atrial fibrillation, n (%) | 7 (33.3%) | 16 (14.7%) | **0.041** |
| Perioperative, n (%) | 8 (38.1%) | 13 (61.9%) | 1 |
| AIS* nadir, median (IQR) | 2 (1, 3) | 3 (2, 4) | 0.11 |

\*American Spinal Injury Association impairment scale

## References

1. Zalewski NL, Flanagan EP, Keegan BM. Evaluation of idiopathic transverse myelitis revealing specific myelopathy diagnoses. Neurology. 2018;90(2):e96–e102.

2. Rigney L, Cappelen-Smith C, Sebire D, Beran RG, Cordato D. Nontraumatic spinal cord ischaemic syndrome. Journal of Clinical Neuroscience: Official Journal of the Neurosurgical Society of Australasia. 2015;22(10):1544–1549.

3. Barreras P, Fitzgerald KC, Mealy MA, Jimenez JA, Becker D, Newsome SD, Levy M, Gailloud P, Pardo CA. Clinical biomarkers differentiate myelitis from vascular and other causes of myelopathy. Neurology. 2018;90(1):e12–e21.

4. Bracken MB, Shepard MJ, Holford TR, Leo-Summers L, Aldrich EF, Fazl M, Fehlings M, Herr DL, Hitchon PW, Marshall LF, Nockels RP, Pascale V, Perot PL, Piepmeier J, Sonntag VK, et al. Administration of methylprednisolone for 24 or 48 hours or tirilazad mesylate for 48 hours in the treatment of acute spinal cord injury. Results of the Third National Acute Spinal Cord Injury Randomized Controlled Trial. National Acute Spinal Cord Injury Study. JAMA. 1997;277(20):1597–1604.

5. Hnath JC, Mehta M, Taggert JB, Sternbach Y, Roddy SP, Kreienberg PB, Ozsvath KJ, Chang BB, Shah DM, Darling RC. Strategies to improve spinal cord ischemia in endovascular thoracic aortic repair: Outcomes of a prospective cerebrospinal fluid drainage protocol. Journal of Vascular Surgery. 2008;48(4):836–840.

6. Caton MT, Huff JS. Spinal Cord Ischemia. In: StatPearls. Treasure Island (FL): StatPearls Publishing; 2025.

7. Anon. SPINAL CORD INFARCTION. CONTINUUM: Lifelong Learning in Neurology. 2005;11(3):87.

8. Sugiura J, Oshima H, Abe T, Narita Y, Araki Y, Fujimoto K, Mutsuga M, Usui A. The efficacy and risk of cerebrospinal fluid drainage for thoracoabdominal aortic aneurysm repair: a retrospective observational comparison between drainage and non-drainage†. Interactive CardioVascular and Thoracic Surgery. 2017:ivw436.

9. Epstein NE. Cerebrospinal fluid drains reduce risk of spinal cord injury for thoracic/thoracoabdominal aneurysm surgery: A review. Surgical Neurology International. 2018;9:48.

10. Coselli JS, LeMaire SA, Köksoy C, Schmittling ZC, Curling PE. Cerebrospinal fluid drainage reduces paraplegia after thoracoabdominal aortic aneurysm repair: Results of a randomized clinical trial. Journal of Vascular Surgery. 2002;35(4):631–639.

11. Strohm T, John S, Hussain M. Cerebrospinal Fluid Drainage for Acute Spinal Cord Infarction (P1.301). Neurology. 2017;88(16_supplement):P1.301.

12. Fehlings MG, Wilson JR, Tetreault LA, Aarabi B, Anderson P, Arnold PM, Brodke DS, Burns AS, Chiba K, Dettori JR, Furlan JC, Hawryluk G, Holly LT, Howley S, Jeji T, et al. A Clinical Practice Guideline for the Management of Patients With Acute Spinal Cord Injury: Recommendations on the Use of Methylprednisolone Sodium Succinate. Global Spine Journal. 2017;7(3 Suppl):203S–211S.

13. Sultan I, Lamba N, Liew A, Doung P, Tewarie I, Amamoo JJ, Gannu L, Chawla S, Doucette J, Cerecedo-Lopez CD, Papatheodorou S, Tafel I, Aglio LS, Smith TR, Zaidi H, et al. The safety and efficacy of steroid treatment for acute spinal cord injury: A Systematic Review and meta-analysis. Heliyon. 2020;6(2):e03414.

14. Liu Z, Yang Y, He L, Pang M, Luo C, Liu B, Rong L. High-dose methylprednisolone for acute traumatic spinal cord injury: A meta-analysis. Neurology. 2019;93(9):e841–e850.

15. Menacho ST, Floyd C. Current practices and goals for mean arterial pressure and spinal cord perfusion pressure in acute traumatic spinal cord injury: Defining the gaps in knowledge. The Journal of Spinal Cord Medicine. 44(3):350–356.

16. Saadeh YS, Smith BW, Joseph JR, Jaffer SY, Buckingham MJ, Oppenlander ME, Szerlip NJ, Park P. The impact of blood pressure management after spinal cord injury: a systematic review of the literature. 2017.

17. Martin ND, Kepler C, Zubair M, Sayadipour A, Cohen M, Weinstein M. Increased mean arterial pressure goals after spinal cord injury and functional outcome. Journal of Emergencies, Trauma, and Shock. 2015;8(2):94–98.

18. Kepler CK, Schroeder GD, Martin ND, Vaccaro AR, Cohen M, Weinstein MS. The effect of preexisting hypertension on early neurologic results of patients with an acute spinal cord injury. Spinal Cord. 2015;53(10):763–766.

19. Inoue T, Manley GT, Patel N, Whetstone WD. Medical and surgical management after spinal cord injury: vasopressor usage, early surgerys, and complications. Journal of Neurotrauma. 2014;31(3):284–291.

20. Readdy WJ, Whetstone WD, Ferguson AR, Talbott JF, Inoue T, Saigal R, Bresnahan JC, Beattie MS, Pan JZ, Manley GT, Dhall SS. Complications and outcomes of vasopressor usage in acute traumatic central cord syndrome. Journal of Neurosurgery. Spine. 2015;23(5):574–580.

21. Levi L, Wolf A, Belzberg H. Hemodynamic parameters in patients with acute cervical cord trauma: description, intervention, and prediction of outcome. Neurosurgery. 1993;33(6):1007–1016; discussion 1016-1017.

22. Sajdeya R, Yanez ND, Kampp M, Goodman MD, Zonies D, Togioka B, Nunn A, Winfield RD, Martin ND, Kohli A, Huynh TT, Okonkwo DO, Poblete RA, Gilmore EJ, Chesnut RM, et al. Early Blood Pressure Targets in Acute Spinal Cord Injury: A Randomized Clinical Trial. JAMA Network Open. 2025;8(9):e2525364.

23. Zalewski NL, Rabinstein AA, Krecke KN, Brown RD, Wijdicks EFM, Weinshenker BG, Kaufmann TJ, Morris JM, Aksamit AJ, Bartleson JD, Lanzino G, Blessing MM, Flanagan EP. Characteristics of Spontaneous Spinal Cord Infarction and Proposed Diagnostic Criteria. JAMA Neurology. 2019;76(1):56–63.

24. Priebe MM, Waring WP. The interobserver reliability of the revised American Spinal Injury Association standards for neurological classification of spinal injury patients. American Journal of Physical Medicine & Rehabilitation. 1991;70(5):268–270.

25. Rankin J. Cerebral Vascular Accidents in Patients over the Age of 60: II. Prognosis. Scottish Medical Journal. 1957;2(5):200–215.

26. Association JO. Scoring system (17-2) for cervical myelopathy. J Jpn Orthop Assoc. 1994;68:490–503.

27. de Seze J, Stojkovic T, Breteau G, Lucas C, Michon-Pasturel U, Gauvrit J-Y, Hachulla E, Mounier-Vehier F, Pruvo J-P, Leys D, Destée A, Hatron P-Y, Vermersch P. Acute myelopathies: Clinical, laboratory and outcome profiles in 79 cases. Brain. 2001;124(8):1509–1521.

28. Salvador de la Barrera S, Barca-Buyo A, Montoto-Marqués A, Ferreiro-Velasco ME, Cidoncha-Dans M, Rodriguez-Sotillo A. Spinal cord infarction: prognosis and recovery in a series of 36 patients. Spinal Cord. 2001;39(10):520–525.

29. Stenimahitis V, Fletcher-Sandersjöö A, El-Hajj VG, Hultling C, Andersson M, Sveinsson O, Elmi-Terander A, Edström E. Long-term Outcomes After Periprocedural and Spontaneous Spinal Cord Infarctions. Neurology. 2023;101(2):e114–e124.

30. Robertson CE, Brown RD, Wijdicks EFM, Rabinstein AA. Recovery after spinal cord infarcts. Neurology. 2012;78(2):114–121.

31. Gharios M, Stenimahitis V, El-Hajj VG, Mahdi OA, Fletcher-Sandersjöö A, Jabbour P, Andersson M, Hultling C, Elmi-Terander A, Edström E. Spontaneous spinal cord infarction: a systematic review. BMJ Neurology Open. 2024;6(1).

